# Service Penetration, Acceptability, Appropriateness, and Fidelity of a Multicomponent Navigation Strategy for HIV-associated Kaposi’s Sarcoma: Mixed Methods Evaluation

**DOI:** 10.1101/2025.04.21.25326038

**Authors:** Sigrid Collier, Merridy Grant, Aggrey Semeere, Helen Byakwaga, Miriam Laker-Oketta, Linda Chemtai, Celestine Lagat, Jolie Phan, Kevin Ng, Anjuli D Wagner, Ingrid V Bassett, Toby Maurer, Jeffrey Martin, Samson Kiprono, Esther E Freeman

**Affiliations:** University of Washington, Seattle, WA USA; Western Australian Centre for Rural Health, University of Western, Australia; Infectious Disease Institute, Kampala, Uganda; Academic Model Providing Access to Healthcare (AMPATH), Eldoret, Kenya; Fred Hutchinson Cancer Center, Seattle, Washington; Massachusetts General Hospital, Harvard Medical School, Boston, MA USA; Indiana University, Indianapolis, IN USA; University of California San Francisco, San Francisco, California USA; Moi University, School of Medicine, Department of Internal Medicine, Eldoret, Kenya

## Abstract

Kaposi’s sarcoma (KS) remains common in sub-Saharan Africa and despite persistently high mortality less than 50% of people with advanced-stage KS with an indication for chemotherapy currently receive it in western Kenya. To address this, a tailored multi-component navigation strategy including physical navigation and care coordination, peer mentorship, education, assistance with health insurance, a health insurance stipend, and transportation stipends was implemented within AMPATH healthcare network in western Kenya in 2021. This study evaluates service penetration (engagement), acceptability, appropriateness, and fidelity to the multi-component navigation strategy.

We used a convergent mixed methods approach using Proctor et al.’s framework for implementation outcomes. We enrolled all adults with newly diagnosed HIV-associated KS from 2021 to 2024. Quantitative data included structured questionnaires, CD4+ T cell count, and navigation activity logs. Scores for acceptability and appropriateness questionnaires ranged from 4 to 20, with 20 representing high levels. In-depth interviews were also conducted among people with HIV-associated KS, healthcare workers, and navigation team members. Descriptive statistics were used for measures of service penetration (engagement), acceptability, appropriateness, and fidelity. Framework analysis was used for in-depth interviews.

Among the 124 Adults with HIV associated KS eligible to participate, service penetration was 74% within 90 days after KS diagnosis. The median acceptability score among people with HIV-associated KS was 20 (Range: 19, 20) and appropriateness was 20 (Range 20, 20). Fidelity to at least one component was 87% (N=80), and no participant experienced all 6 components. Fidelity was 2.2% (N=2) for the transportation stipends (7 total) and 28% (N=26) for the health insurance enrollment stipend. During in-depth interviews, patients and healthcare workers described high levels of acceptability and appropriateness of the navigation strategy. Patients described experiences of variability in navigation strategy components, and healthcare worker and navigation team members described how financial constraints and time constraints contributed to variability in fidelity.

A multi-component navigation strategy designed to improve chemotherapy engagement for HIV-associated KS was both acceptable and appropriate. Fidelity was variable with low fidelity to financial components, suggesting areas for future adaptation to ensure sustainability and context appropriateness during integration into the health system and future scale-up.

## Introduction

Despite increasing antiretroviral therapy (ART) use, Kaposi’s sarcoma (KS) remains one of the most common HIV-associated malignancies in sub-Saharan Africa, accounting for over 70% of the global incidence of KS.^1^ KS is often diagnosed at an advanced disease stage in this setting, and mortality is estimated to be 45% within two years after KS diagnosis.^2^ Using ART in combination with chemotherapy has been shown to reduce morbidity and mortality among people with advanced-stage KS, improving KS response rates by approximately 20-40% compared to ART alone.^3–6^ Although chemotherapy can significantly improve clinical outcomes, less than 50% of people with advanced-stage KS with an indication for chemotherapy are receiving it in western Kenya.^7^

In our prior work, we identified barriers to chemotherapy initiation and adherence for KS in western Kenya which include a lack of financial means, challenges with the distance to healthcare facility, intrapersonal barriers (e.g., fear and hopelessness), and lack of education about KS and chemotherapy.^8^ Our team worked to design a multi-component strategy that was tailored to address these barriers to KS care in western Kenya using intervention mapping.^9^ The strategy was built around the concept of patient navigation, which is an evidence-based, community-oriented strategy to improve disparities in cancer care.^10^ Our tailored multi-component navigation strategy included the following components: physical navigation and care coordination, peer mentorship, education using two videos about KS diagnosis and KS treatment, assistance with health insurance, a health insurance stipend, and a transportation stipend for the first oncology visit and all chemotherapy visits.^9^

While navigation is an established strategy for addressing barriers to care for vulnerable populations and improve access to care for cancer and other chronic conditions, there have been few studies evaluating navigation strategies for HIV-associated malignancies in LMICs. ^11–13^ In this study, we describe the results of our convergent mixed-methods approach to evaluate service penetration (engagement), acceptability, appropriateness, and fidelity to a multi-component navigation strategy for patients with newly diagnosed HIV-related KS in western Kenya.

## Methods

### Overall Study Design

We prospectively enrolled all adults with newly diagnosed HIV-associated KS within a single hospital system in western Kenya, where they had newly implemented a multi-component navigation strategy. To understand service penetration (engagement), acceptability, appropriateness, and fidelity to this multi-component navigation strategy we used a convergent mixed-methods approach combining and triangulating quantitative and qualitative data. We also enrolled healthcare workers and navigation team members as part of a qualitative approach to understanding the implementation of the multi-component navigation strategy.

### Population

This study took place at the Academic Model Providing Access to Healthcare (AMPATH) in Eldoret, Kenya. The KS Center for Excellence at AMPATH implemented a multicomponent patient navigation strategy in 2021, to complement existing dermatology and oncology services available within the AMPATH healthcare system, including diagnostic biopsy and treatment services for KS. AMPATH KS Center of Excellence invites any clients with newly diagnosed HIV-associated KS to participate in the multicomponent patient navigation strategy.^14^ This study prospectively enrolls all adults with newly diagnosed HIV-associated KS who are able to provide informed consent. This includes adults who have either biopsy confirmed KS or are diagnosed with KS on clinical grounds alone when a biopsy is unsafe (e.g., some oral lesions and conjunctival lesions) and those with both a new and/or existing HIV diagnosis. All patients who are diagnosed with KS are screened using clinical evaluation, AIDS Clinical Trials Group Oncology Committee (ACTG) staging criteria, and WHO staging criteria for KS.^15, 16^ We also enrolled healthcare workers, including oncology physicians, clinical officers, nurses, social workers, patient navigators, peer mentors, and health insurance officers, who are involved in the multicomponent patient navigation strategy and/or the care of patients with KS to understand the impact of the implementation strategy on routine clinical care.

### Navigation Strategy

The navigation strategy was implemented by the KS Center for Excellence in September of 2021. Peer and patient navigators were hired by the KS Center of Excellence and were trained using adapted and abbreviated training modules from the “Oncology Patient Navigator Training: The Fundamentals” course from the George Washington University School of Medicine and Health Sciences. This included hands-on practice sessions with case-based scenarios to practice using patient-centered communication techniques. Oncology staff also gave a lecture on cancer care and KS specific treatment that was specific to the AMPATH healthcare system. As part of the training the peer and patient navigator were introduced to clinic staff in the Oncology clinic and other clinical staff involved in KS care (pharmacy, social work, etc.).

The multicomponent navigation strategy was comprised of 6 components, and each of the components had sub-components.

#### Physical navigation and care coordination

The responsibilities of the patient navigator included assistance arranging transportation to oncology and chemotherapy visits, meeting clients on arrival to the health center and physically guiding the clients to their first oncology and first chemotherapy appointments, oncology and chemotherapy visit reminders, guiding clients to health insurance enrollment assistance, and connection to other social services based on each patient’s needs.

#### Peer mentorship

Clients were introduced to the peer mentor by the KS Center of Excellence team at the time of KS diagnosis. The peer mentor is a KS survivor from the same region in western Kenya.

#### Education

The education component was comprised of two educational videos available in Kiswahili (the most common local language). The first educational video was a general video about KS, focusing on the etiology of KS, the natural disease course of KS, and KS diagnostic procedures. The second educational video focused on the treatment of KS, including ART for people with HIV-associated KS, the treatment options for KS, detailed explanations of chemotherapy regimens for KS, and potential side effects from chemotherapy.

#### Assistance with enrollment in health insurance

Clients who were not registered were connected by patient navigators to health insurance officers who assisted them in registering for the Kenyan health insurance, then called the National Health Insurance Fund.

#### Transportation stipend

The strategy included a stipend for all participants to assist with the cost of their transportation for KS-related oncology care and treatment until the completion of their prescribed treatment course.

#### Health insurance stipend

The navigation strategy was designed to pay for the entire first 1-2 years of health insurance for all clients. At the time of the study the national health insurance program in Kenya was called the National Health Insurance Fund (NHIF). Under the NHIF system, even if you were enrolled and up-to-date on payments, there was a required co-pay of either 1 or 2 years of NHIF payments in order to be eligible for chemotherapy.

### Measurements

#### Quantitative

All adults with newly diagnosed HIV-associated KS completed a physical exam, blood draw (including viral load and CD4+ T-Cell count), and a set of structured questionnaires including demographics, HIV and HIV treatment history, and symptoms associated with advanced stage disease (edema, hemoptysis, etc.) at enrollment.

#### Service Penetration and Fidelity to the Implementation Strategy

Service Penetration was defined to be the number of people with HIV-associated KS who had contact with either a patient navigator or a peer mentor within 90 days after KS diagnosis, among all people with HIV-associated KS in the parent study who were eligible for navigation. We defined the date of KS diagnosis as the date when patients were notified of KS biopsy results when that was available and the date of KS biopsy when there was no available record of biopsy result notification.

KS Center of Excellence Staff, including the patient navigation and peer mentor, collected detailed logs of navigation strategy activities, after each patient encounter, whether in-person or via telephone. This included the date of the encounter, what was discussed, and what navigation activities occurred as part of the patient encounter. These detailed logs were used to assess important process outcomes including dose and fidelity to the implementation strategy.

Fidelity was defined for each of the components of the multi-component navigation strategy as follows:

##### Physical navigation and care coordination

Fidelity to physical navigation was defined as receiving all three of the following components: physical navigation to visits (at least twice), reminders about health insurance registration among people with HIV-associated KS not yet registered (at least once), reminders about clinic visits (at least 7 occurrences, one for the first oncology consultation and 6 additional reminders for the 6 chemotherapy treatments which are standard for the current first line chemotherapy). We expected patient navigators to interact with patients a total of 9 times (2 encounters of physical navigation and 7 telephone reminders about oncology and chemotherapy appointments).

We intended to include connecting people with HIV-associated KS to services (at least once) as a component of fidelity, but we were not able to assess this because at the time of designing the strategy and fidelity measures, we did not clearly define what services were included in this definition.

##### Peer Mentorship

Fidelity was defined as a peer speaking to people with HIV-associated KS on the phone or in-person on at least 7 separate occasions including the following at least once: their experience (at least once), guidance on next steps (at least once), and encouragement to continue KS treatment (at least once). We expected the peer mentor to interact with the patient on a total of 7 occasions (after the first oncology visit and after each of the chemotherapy visits).

While we originally intended to include the peer referring to the patient navigator when services were needed (at least once), we were not able to assess this because we did not clearly define and collect data to identify people with HIV-associated KS who needed services.

##### Educational Videos

Fidelity was defined as watching both of the educational videos at least once.

##### Assistance with Enrollment in Health Insurance

Fidelity was defined as receiving at least one reminder about health insurance enrollment.

##### Transportation Stipend

We defined fidelity as receiving a transportation stipend for at least 7 oncology and/or chemotherapy visits.

##### Health Insurance Stipend

Fidelity was defined as receiving at least one NHIF stipend.

#### Acceptability and Appropriateness of the Implementation Strategy

We adapted and field tested the Acceptability of Intervention Measure (AIM) and the Intervention Appropriateness Measure (IAM) measures.^17^ Among all people with HIV-associated KS who were engaged in the navigation strategy (see definition of service penetration above), at 12-, 24-, and 48-weeks, we collected AIM and IAM measures for the multicomponent navigation strategy overall, as well as the KS diagnosis video, KS treatment video, patient navigator, and peer mentor. Our original design included measuring the acceptability and appropriateness of the health insurance stipend and transportation stipend, but after feedback that these items felt redundant and tedious we eliminated these items to reduce the question burden. The AIM and IAM measures are composed of 4 items that are measured on a Likert scale between 1 and 5, with 5 representing the highest levels of acceptability and/or appropriateness.

#### Qualitative

In-depth patient interviews were conducted by experienced Kenyan qualitative interviewers in the patient’s preferred language with a purposively selected subset of 24 people with HIV-associated KS, selected to represent distinct age groups, tribes, and sex with a lived experience of the navigation strategy. The research team attempted to recruit patients who chose not to participate in the navigation strategy but were unable to do so as many declined invitations because of travel time and/or inconvenience. In-depth interviews were carried out at time points at least 1 month and up to 3 months following KS diagnosis to explore experiences of the navigation strategy, focusing on acceptability, appropriateness and utility of its different components and recommendations for improvement.

In-depth interviews were also carried out by trained qualitative interviewers with 14 health care workers and 5 members of the navigation team in the Spring of 2023 at the hospital where the navigation strategy was implemented. Health care workers were purposively selected based on their role along the cascade of care for KS patients and included waiting room staff, oncology-based clinical officers, nurses, managers, patient attendants and record officers, hospital dermatologists, hospital social workers, pharmacists and hospital health insurance staff involved in preparing and authorizing health insurance claims. All navigation team members participated in in-depth interviews. Interviews explored knowledge of KS, attitudes and experiences of the strategy, including its acceptability, appropriateness, utility and recommendations, and for navigation team interviews they also explored the barriers and enablers to navigation implementation. Interviews were audio recorded and were approximately 60-minutes in length.

### Analysis

#### Quantitative

##### Service Penetration and Fidelity to the Implementation Strategy

We calculated proportions for service penetration. We also calculated proportions for fidelity, including fidelity to all 6 components, and fidelity to each of the components and sub-components. We also calculated medians and interquartile ranges for the doses of patient navigation, peer mentorship, and educational videos.

##### Acceptability and Appropriateness of the Implementation Strategy

We calculated summative scores for each of the Acceptability of Intervention Measure (AIM) and the Intervention Appropriateness Measure (IAM) measures.^17^ We calculated the median and interquartile range for AIM and IAM for the multicomponent navigation strategy overall, as well as the KS Diagnosis Video, KS Treatment Video, Patient Navigator, and Peer Mentor. In exploratory analyses we also calculated the median, minimum, and maximum AIM and IAM score across all time points for people with HIV-associated KS who answered the questionnaire at least once during the study period. We then calculated the median of medians, minimum of minimums, and maximum of maximums for all participants with HIV-associated KS across all time periods.

#### Qualitative

The recorded in-depth interviews were transcribed and translated from the language in which the interview was performed (Swahili, English, or other local dialect) into English by trained Kenyan research assistants. Framework Analysis was conducted with the aid of NVivo software.^18^ Three qualitative analysts (SC, JP, MG), developed and refined the thematic structure, completed coding and analysis of the patient data. For the health care worker and navigation team interviews, two qualitative analysts (SC and MG) performed the same process. For all interviews, the core components of the navigation strategy informed the overarching *a priori* themes, while allowing for themes to emerge inductively per the framework analysis method.^18^ For health care worker and navigation team interviews, analysts added an additional theme of fidelity guided by the quantitative findings.

#### Triangulation

Triangulation was carried out by using different data collection methods (in-depth interviews, questionnaire data) and accessing information from diverse sources (patients, health care workers, navigation strategy team members), to gain a comprehensive understanding of the navigation strategy.

## Results

### Characteristics of the Study Population

During the study period, there were 124 adults (18+) with newly diagnosed HIV-associated KS within the AMPATH system who were eligible to participate in the multi-component navigation strategy. Among these 124 adults, most were men (61%) and the median age was 38 (Table 1). Most people with HIV-associated KS had completed primary education (71.8%). The median income per month was 5000 kSh per month (approximately 39 USD). The median CD4+ T-Cell count was 160 cells/mm.^3^

**Table 1:**
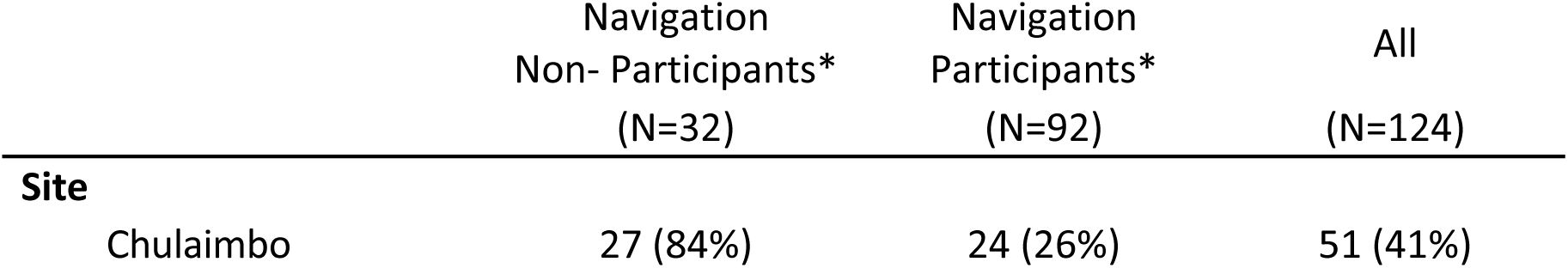
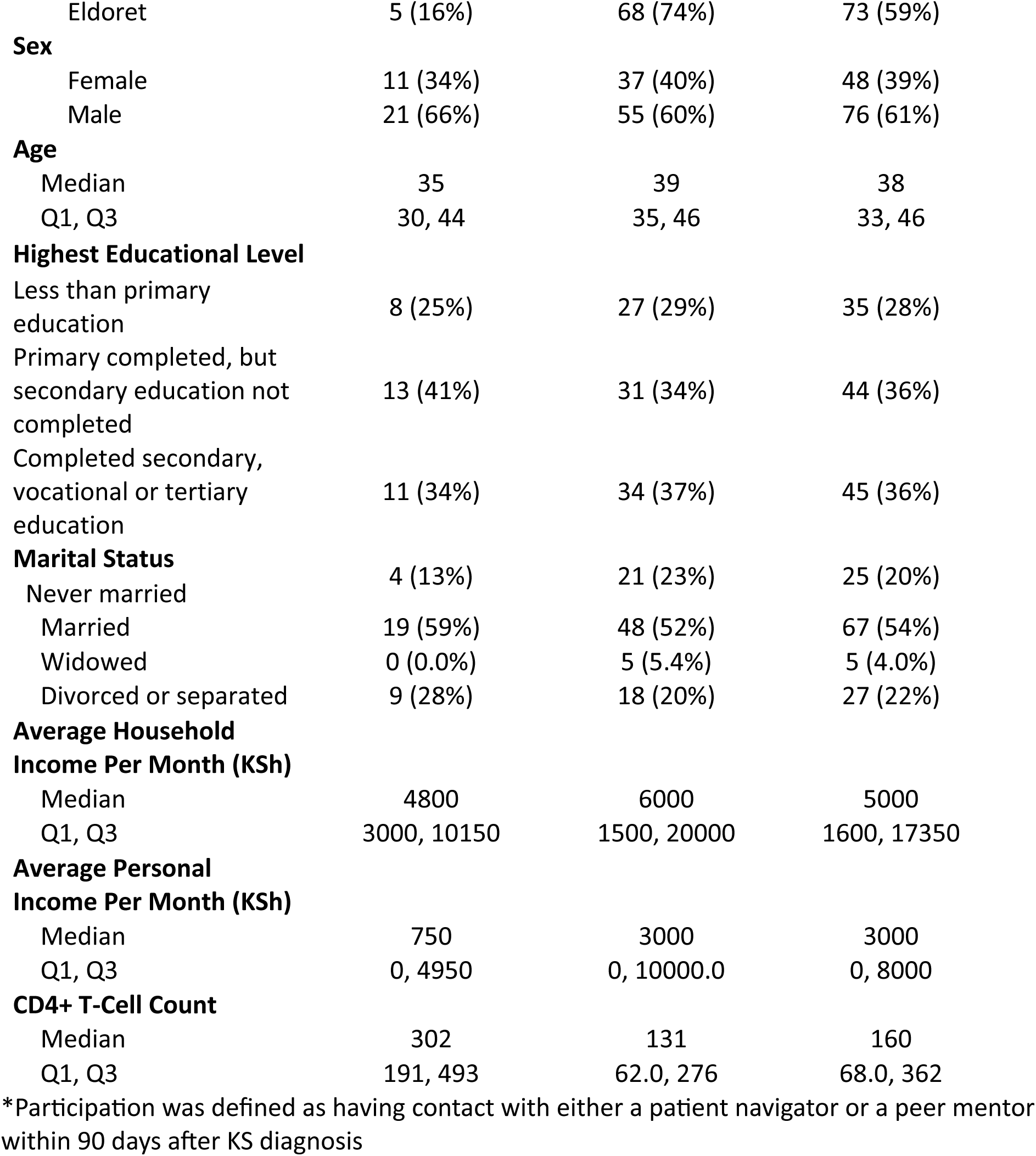
Demographics by Service Penetration.

|  | Navigation<br>Non- Participants*<br>(N=32) | Navigation<br>Participants*<br>(N=92) | All<br>(N=124) |
| --- | --- | --- | --- |
| **Site** |  |  |  |
| Chulaimbo | 27 (84%) | 24 (26%) | 51 (41%) |
| Eldoret | 5 (16%) | 68 (74%) | 73 (59%) |
| **Sex** |  |  |  |
| Female | 11 (34%) | 37 (40%) | 48 (39%) |
| Male | 21 (66%) | 55 (60%) | 76 (61%) |
| **Age** |  |  |  |
| Median | 35 | 39 | 38 |
| Q1, Q3 | 30, 44 | 35, 46 | 33, 46 |
| **Highest Educational Level** |  |  |  |
| Less than primary education | 8 (25%) | 27 (29%) | 35 (28%) |
| Primary completed, but secondary education not completed | 13 (41%) | 31 (34%) | 44 (36%) |
| Completed secondary, vocational or tertiary education | 11 (34%) | 34 (37%) | 45 (36%) |
| **Marital Status** |  |  |  |
| Never married | 4 (13%) | 21 (23%) | 25 (20%) |
| Married | 19 (59%) | 48 (52%) | 67 (54%) |
| Widowed | 0 (0.0%) | 5 (5.4%) | 5 (4.0%) |
| Divorced or separated | 9 (28%) | 18 (20%) | 27 (22%) |
| **Average Household Income Per Month (KSh)** |  |  |  |
| Median | 4800 | 6000 | 5000 |
| Q1, Q3 | 3000, 10150 | 1500, 20000 | 1600, 17350 |
| **Average Personal Income Per Month (KSh)** |  |  |  |
| Median | 750 | 3000 | 3000 |
| Q1, Q3 | 0, 4950 | 0, 10000.0 | 0, 8000 |
| **CD4+ T-Cell Count** |  |  |  |
| Median | 302 | 131 | 160 |
| Q1, Q3 | 191, 493 | 62.0, 276 | 68.0, 362 |

\*Participation was defined as having contact with either a patient navigator or a peer mentor within 90 days after KS diagnosis

### Service Penetration and Dose of the Navigation Strategy, Quantitative

Among the clients who qualified for participation in the navigation strategy, 74.2% (n=92) had contact with either a patient navigator or a peer mentor within 90 days after KS diagnosis (Table 1).

### Dose

People with HIV-associated KS had a median of 20.5 interactions with the patient navigator, instead of the expected 9. In contrast, people with HIV-associated KS had a median of 5 interactions with the peer, instead of the expected 7. The strategy was designed for people with HIV-associated KS to watch 2 educational videos, and they watched a median of 2 educational videos.

**Table 2:**

| Outcome Measures | Expected | Outcome (N=124) |
| --- | --- | --- |
| **Service Penetration** | 100% | 74% (92) |
| **Dose** |  |  |
| **Patient Navigation** | 9 interactions | 20.5 |
| **Peer Mentorship** | 7 interactions | 5.0 |
| **Educational Videos** | 2 videos watched | 2.0 |

### Acceptability and Appropriateness of the Navigation Strategy, Quantitative

Among those who participated in the strategy, a total of 83 (96%) responded to questions about the acceptability and appropriateness of the multicomponent navigation strategy and its sub-components at least once during the study period. The median and interquartile range for acceptability and appropriateness scores at 12-, 24-, and 48-weeks are reported in table 3. The median acceptability scores across all time points among people with HIV-associated KS who completed the acceptability and appropriateness questionnaires at least once was 20 with a range from 19 to 20 for acceptability and 20 with a range of 20 to 20 for appropriateness (Table 3).

**Table 3:** Acceptability and Appropriateness of Navigation Strategy among People with HIV-Associated KS.

| **Acceptability** |  |
| --- | --- |
|  | **N=83**<br>**Median Across all Timepoints**<br>**(Min, Max)** |
| **Multicomponent Navigation Strategy Overall** | **20 (19, 20)** |
| Physical Navigation | 20 (16, 20) |
| Peer Mentor | 20 (16, 20) |
| Education | Diagnosis Video: 20 (4,20)<br>Treatment Video: 20 (16, 20) |
| Transportation Stipend | _____ |
| Health Insurance Enrollment Stipend | _____ |
| **Appropriateness** |  |
|  | **N=83**<br>**Median (Min, Max)** |
| **Multicomponent Navigation Strategy Overall** | **20 (20, 20)** |
| Physical Navigation | 20 (16, 20) |
| Peer Mentor | 20 (16, 20) |
| Education | Diagnosis Video: 20 (4,20)<br>Treatment Video: 20 (16, 20) |
| Transportation Stipend | _____ |
| Health Insurance Enrollment Stipend | _____ |

### Acceptability and Appropriateness of the Navigation Strategy, Qualitative

In interviews, all patients (n=24) found the navigation strategy to be acceptable and spoke highly of the navigation strategy. One participant shared “***Personally it’s very attractive, supportive and has helped me a lot. I haven’t found anywhere else with this kind of support.” (ELD 1009, Patient Interview).*** Additionally, all health care workers (n=14) found the navigation strategy to be acceptable and reported that the navigation strategy added value to KS patients and the oncology program in the facility:

> ***…the navigation program is actually what the KS patients need, and they needed this program for a long time… because these are the challenges we had initially …so you could work as clinicians and at the same time you become a navigator, so you used to waste a lot of time navigating, going around assisting these patients (ELDPRO1012, Health care worker Interview)***

All patients and health care workers reported that the navigation strategy was appropriate to their context and treatment process. Reasons for this included that the components of the navigation strategy saved lives and helped patients to recover:

> ***First thing the program has helped me in geĖng treatment faster because of these peers…navigators… if I hadn’t met with them, I wouldn’t know where to start…because you know this disease it actually needs support [it] may be emotional support and [it] may be financial support because you can come here and then the finances [overwhelm you]…you find that it can make you say that let me just die. So, navigation is good you have assisted so many and I can say it has helped me a lot… because it is always good if you are given treatment and you understand how the treatment is progressing and at the same time you also understand how you are doing (CHU1018, Patient Interview)***

Health care workers reported that the KS navigators streamlined the cascade of care, positively impacting turnaround times for results and uptake of chemotherapy:

> ***So for me, the first one, which is working very well [due to the navigation program], is the turnaround time for the patient to get the results… So the patient really doesn’t have to wait…for like two weeks…The second thing that I’m seeing that is working very well, is the uptake for chemotherapy…when these patients go through the sessions with the peer mentors, especially when they are educated on the chemo, they look at the videos and all that. I think it [uptake of chemo] improves… because we used to have so many patients who declined chemo, because of fear…culturally, if you look at our people, they always say “no! no!” they say that “we are given chemo then it’s like a death sentence.” (ELDPRO1008, Health care worker Interview)***

### Implementation Fidelity and Patient Perspectives on the Multicomponent Navigation Strategy

#### Overview, Quantitative

Table 4 outlines the fidelity to each of the components and sub-components. Of the 92 clients who met the criteria for service penetration (engagement) in the multicomponent navigation strategy, 87% (N=80) received at least one of all six major components exactly as it was designed. None of the participants received all 6 components exactly as they were designed.

**Table 4:** Fidelity to Navigation Strategy Components and Sub-Components.

| Component | Fidelity (N=92) |
| --- | --- |
|  | % (N) |
| **Physical Navigation** | 47% (43) |
| 1) Physically navigate to visits at least once | 95% (87) |
| 2) Reminder about health insurance registration at least once* | 47% (7) |
| 3) Reminder about appointments at least 7 occurrences | 47% (43) |
| **Peer Mentor** | 39% (36) |
| Speak to person with KS on the phone or in-person on at least 7 separate occasions including the following at least once. | 39% (36) |
| 1) their experience | 43% (40) |
| 2) guidance on next steps | 80% (74) |
| 3) encouragement to continue Ks treatment | 84% (77) |
| **Education** | 54% (50) |
| 1) Video about KS etiology and disease course at least once | 62% (57) |
| 2) Video about KS treatment at least once | 61% (56) |
| **Transportation Stipend** |  |
| Proportion of people with HIV-associated KS who received at least one transportation stipend. | 59% (54) |
| Proportion of people with HIV-associated KS who received a stipend to offset the cost of transportation to a total of 7 oncology and/or chemotherapy visits. | 2.2% (2) |
| **Health Insurance Enrollment Assistance\*** | 78% (72) |
| **Health Insurance Enrollment Stipend** | 28% (26) |

\*Among people with HIV-associated KS who were not previously registered for NHIF (N=15)

#### Overview, Qualitative

In both patient and health care worker interviews, participants were able to describe the navigation strategy in detail including help with NHIF paperwork and costs, accommodation, transport fare, physical navigation of the hospital departments and systems, treatment reminders and peer navigation.

> ***When I started they [Patient Navigator and Peer Mentor] were with me throughout even when I started the first clinic, because I didn’t know where to start from, when I came and I was given my file, we were together with her [Patient Navigator], she directed me to go to NHIF, she took me to the triage, from there to the laboratory where they tested my blood, she still took me to all these places until finally I was seen by the doctor. From there, she took my file and we went to the Pharmacy, from the Pharmacy back to NHIF until we got to Chemo, and medicine was administered. So they have been very helpful to me. Because even on my revisits, how they advised me, how they directed me, I can now comfortably go through the clinic (ELD1092, Patient Interview)***

##### Physical navigation and care coordination, Quantitative

Of the people with HIV-associated KS who participated in navigation, 53% (n=49) received all 3 components of physical navigation (Table 4). Most people with HIV-associated KS were physically navigated during at least one clinic visit, 95% (N=87).

##### Physical navigation and care coordination, Qualitative

In interviews, physical navigation was highly valued by all patients (n=24) and health care workers (n=14). People with HIV-associated KS who lived far away and/or were visiting the hospital for the first time described the particular significance for those situations. Many patients referred to feeling fearful on their first visit and described immense relief once meeting the navigator as the hospital systems were confusing and intimidating:

> ***Patient Navigator [The navigator] has been very helpful because it was my first time here, and I didn’t know where to start or finish, but she held my hand, because I was sick, I was confused, I didn’t know what to do I. (ELD1092, Patient Interview)***

> ***I’ve seen them come with patients, I don’t know where they get them from, patients with suspicious lesions, they come with them, they assist them in doing biopsy tests. They do follow up, when the histology results are in, they call the patients, the patients come back. And now in the clinic when we are seeing these patients, they actually come with them, literally they walk with them to the room, they show them, they introduce them to us, and they give them their privacy… they move out as we see the patients. After making our treatment planning, then they come and also walk with them to the next step, that is the pharmacy. And after pharmacy they sometimes give us feedback, telling us how the patients are faring on at home after treatment, and actually the navigation program…I have liked it, I wish every program or every clinic should have a navigator. (ELDPRO1012, Health care worker Interview)***

Many people with HIV-associated KS described less support after the first visit once they felt more confident navigating the space and were provided with the navigator’s phone number in case they needed assistance:

> ***when I came back because I was with someone else, they wanted to see if I could manage or not, but she was monitoring me, she would come in between intervals and ask where I had reached, and I would say, I am at a certain point, then she would say from here go to a certain point. So after that, by the 3rd visit I had mastered the process. So when she was dealing with newer patients, I was taking care of myself. Up to now I know where to start from and where to finish. (ELD1092, Patient Interview)***

A KS navigation team member explained that as the number of patients increased, the team had to prioritize new patients and high needs (in need of physical or financial help) patients:

> ***So, [in] maintaining the program, I think since the patients are so many at the clinic, just to facilitate the program to give us more navigators, [it] would really assist in this… because some patients feel left out at some point, because we cannot get all of them, so we prioritize on the new patients, and the needy patients that really need the navigators to assist them (RCAPRO7005, KS Navigation Team Interview)***

The high patient load, the intense care needs of KS patients, many of whom were unable to walk unassisted and were without social support, and the need for more navigators was a common theme across interviews (patients, health care workers and KS navigation team):

> ***The challenge…is the number of patients, and some patients are very sick. Like now there was a patient, I can’t remember the name, the patient was…on [a] stretcher. …there are so many patients, not one. So they follow through that [patient] on the stretcher, there [are] others in wheelchairs, so the number of patients is very high. (ELDPRO1006, Health care worker Interview)***

Many patients interviewed reported how much they valued the encouragement from the physical navigator and described an associated sense of hope that they would recover. Patients also described a strong sense of trust and rapport with the physical navigator.

> ***Meeting with her gave me hope. The way she explained to me how the medicine would help me. She would show me how other people were, but they changed and got better That gave me hope that I would be like so-and-so, because I have seen with my eyes what she explained to me. That gave me hope to continue following up with her. (ELD1093, Patient Interview)***

Health care workers acknowledged the encouragement and support navigators demonstrated toward patients throughout the treatment process.

> ***They [navigators] also make patients feel welcomed: and then for the patients to feel they are welcome, to feel they are honored, to feel they are not discriminated and then they are really assisted in that pathway of treatment until the end, and at some point they also follow up on how they are going…after the medication. (ELDPRO1014, Health care worker Interview)***

> ***You know, most of our patients come in with Kaposi sarcoma, they have wounds, they’re smelling and all that, so no one wants to be around them. But now you find a patient navigator is always like working with you, despite your wounds, despite your smell, you know, and yeah, so it really helps the patient… [ELDPRO1013, Health care worker Interview)***

##### Reminders for Chemotherapy, Quantitative

Fewer KS patients received appointment reminders for all 7 oncology and chemotherapy appointments, 47% (N=43). This was reflected in patient and health care worker interviews, which described variable consistency in clinic reminders and logistical challenges, respectively.

##### Reminders for Chemotherapy, Qualitative

In interviews, a few patients spoke of the importance of the reminders, with some noting they allowed time to raise transport money. When asked about patient electronic visit reminders, one of the navigators emphasized that phone calls to remind patients of their treatment days were invaluable in encouraging treatment adherence.

> ***Then the reminder, when you are reminded on Friday, at least you start running up and down looking for bus fare, some of us come from very far and you have to use bus fare which you have to look for. Some of us are not working, so you are forced to sell a tree or call a brother to bail you out… So the reminder also has been very helpful. (ELD1092, Patient interview).***

In interviews, two navigation team members explained that the tracking system for reminders needed to be streamlined as reminders for some patients were sometimes forgotten. The paper-based system was challenging, especially given that some patients changed their treatment days. Both navigation team members suggested an electronic system to track patients.

> ***Sometimes patients are lost. Let’s say we have 100 patients, keeping track for 100 patients just to know the days that they’re supposed to come back to the clinic is a bit hectic, if it [tracking] can be … [electronic] system reminders, not physical reminders, as we do on paper, […](RCAPRO7002, Navigation Team Interview)***

##### Peer Mentorship, Quantitative

Most patients who participated in navigation strategy met with a peer mentor at least once (84%), while only 39% of people with HIV-associated KS received all 3 components of peer mentorship as designed (Table 4). Most patients received encouragement to continue chemotherapy (84%) and guidance around next steps (80%), while the peer shared his or her personal experience with fewer people with HIV-associated KS (43%).

##### Peer Mentorship, Qualitative

In interviews, all people with HIV-associated KS (n=24) described a sense of encouragement and hope from engaging with the peer mentor. Many patients reported that the peer mentor helped them understand KS and KS treatment better. Most patients (n=22) felt that the peer mentor deeply understood their challenges through shared experiences, giving them hope and encouragement for recovery.

> ***The one who has been through the illness, is talking about the real thing. It is not like the one who has not been sick. It is only the one who has been through the pain that understands. The one who has not been through that cannot be told. They have not felt it as it is. (ELD1116, Patient Interview)***

The peer mentor’s role in providing mental health support was invaluable:

> ***First of all, he treated my thoughts. I had a lot of thoughts. But from his pictures and videos, that was treatment. He treated my thoughts so I stopped thinking that I was sick. I started thinking that I was better. That was the first thing he did. (ELD1093, Patient interview)***

> ***Mental support is often more valuable than physical support. I mean, you can give me money, but if I have a negative mindset, that money won’t ultimately help me. I might even consider harming myself, rendering your money useless. Therefore, it’s crucial to prioritize giving [the peer mentor] more time to talk to patients (ELD1009, Patient Interview).***

##### Education, Quantitative

Of the people with HIV-associated KS who engaged with the navigation strategy, 54% (n=50) of people with HIV-associated KS watched both the KS diagnosis and KS treatment videos at least once.

##### Education, Qualitative

One navigation team member explained that not all patients were interested in the education components and just wanted the transport fare:

> ***There is a patient who came and while we were talking he wasn’t cooperating much, you find that you are showing him a video and when his phone rings he picks the call, … but when it comes to the time he needs fare, he wants fare so, you see for such a person his concern is that money, not to be guided. (RCAPRO7003, Navigation Team Member Interview)***

In interviews, almost all patients (n=20) reported that the videos were helpful. They described that the videos gave encouragement and hope that they would heal/recover, helped them understand and accept their condition, and motivated them to initiate treatment.

> ***His video is what gave me the reassurance that he was not lying to me. I saw it with my own eyes, and I saw the person who was next to me. (ELD1093, Patient Interview)***

> ***What touched me most is that you see when I got this disease I felt like I would never live any more, but when I got to watch that video and I saw people geĖng healed I got that hope even me-I will just be healed. (ELD1008, Patient Interview)***

> ***Yes, it is those videos that [the peer mentor] showed me, they are the ones that encouraged me and gave me the most push because in the same video there was a part where someone died having not been treated so that encouraged me and I was like I can continue siĖng there, then I end up not geĖng treatment and then I also die. (ELD 1008, Patient Interview)***

The first video shown, which was focused on KS diagnosis and the natural course of disease, including a scene about the risk of death was adapted from a video developed by the team for a related project in Uganda. In interviews, a few navigation team members suggested that the death scene be edited out of the diagnosis video. Four patients found the first video (which included the death scene) shocking and fear-inducing. Interestingly three out of four of these people with HIV-associated KS still recommended that videos be shown to other patients in the future, while one suggested meeting the peer mentor first.

> ***When I saw it, I just knew I am going to die but [the peer mentor] talked to me, [the navigator] and the doctor also counselled me and when I went home, I was very happy. (ELD1099, Patient Interview)***

##### Assistance with enrollment in health insurance, Quantitative

Most people with HIV-associated KS 78% (n=72) who engaged with the navigation strategy received assistance with the Kenyan national health insurance. Some of the patients (N=15) had not completed the first registration step for health insurance, and among those patients only 47% (N=7) received reminders about health insurance registration.

##### Assistance with enrollment in health insurance, Qualitative

In patient interviews, most participants (n=21) found assistance with health insurance enrollment important for accessing and completing treatment, with three participants not mentioning receiving such support. Patients described the importance of the navigator (and occasionally the peer mentor’s) role in facilitating insurance paperwork, following up with NHIF, and navigating NHIF related challenges. Two people with HIV-associated KS specified that the health insurance assistance was the most important or helpful part of the strategy.

> ***The only part I can say made me… get treatment fast was that part of NHIF, the process of filling the forms and geĖng the approval message, of which [the navigator] was the one helping me so much with that, she helped me and guided me on how to fill that NHIF form as well as following it up at the NHIF offices so that really helped me. (CHU 1018, Patient Interview)***

Almost all health care workers (11/14) acknowledged the important role the navigators played in assistance with health insurance enrollment:

> ***[The navigators] come …up with the NHIF form for pre authorization for a patient, the patient is not there, but they are following up. …at the end of the day, whatever they’re doing, even us as pharmacy, they are making our work easier. You see, because these are patients, he comes to the NHIF card, NHIF form approved, you are happy, you will not turn him away (ELDPRO1011, Health care worker Interview)***

> ***They do assist those patients to fill the insurance forms, because most of our patients, let’s say half of them like a half are illiterate, so they need some assistance, to fill those forms, so these are important issues, to… assist these patients with. (ELDPRO1012, Health care worker Interview)***

##### Health insurance stipend, Quantitative

While most people with HIV-associated KS participating in the navigation strategy received assistance with health insurance enrollment, only 28% (n=26) received a health insurance stipend to help cover the out-of-pocket cost of chemotherapy under the NHIF system.

##### Health insurance stipend, Qualitative

In patient interviews, half of the participants reported that the health insurance stipend was essential for affording treatment. The degree of financial assistance for health insurance varied; some participants received initial support before covering costs themselves, some paid half and others were fully covered. Two people with HIV-associated KS shared that the health insurance stipend was the most important component of the strategy.

> ***What helped me the most was the NHIF payments. That is what opened up my hope for treatment, because if it had not been paid, I would still be saying that I am looking for money. It helped me the most (CHU 1037, Patient Interview)***

Some health care workers reported that the navigation strategy had played a significant role in financially assisting impoverished patients to enroll in health insurance, making their treatment possible.

> ***Kaposi Sarcoma patients…they come from low social economic zones, they are usually poor, and most of them do not have money, and NHIF has really worked well with them, any patient who has NHIF is usually through the care and we’ve had cases where the program has been helping them … as in enroll them to the program so that they can be treated. (PRO1001, Health care worker Interview)***

A KS navigation team member explained that not all patients received a stipend as the team tried to identify patients in need of assistance. They went on to recommend a screening tool to assist with stipend allocation:

> ***We try to look at the needy patients, because sometimes someone has not paid NHIF, but they can afford [it], it’s only that maybe they thought they don’t need the insurance… to pay for the insurance. But once you tell them and they see the need, they can pay. And there are patients who are totally needy and are not able to [pay]. So part of the things I would want to… maybe I would wish that we have is, could we have like a screening tool, just for the, for the I think it’s like socioeconomic status, just screen so that you can know who qualifies like to be needy, because my assessment and the other person’s assessment might not be the same. (RCAPRO7002, Navigation Team Member Interview)***

##### Transportation stipend, Quantitative

While many people with HIV-associated KS received at least one transportation stipend (59%, N=54), among people with HIV-associated KS who engaged with the navigation strategy only 2.2% (n=2) received a transportation stipend for all 7 oncology and chemotherapy visits.

##### Transportation stipend, Qualitative

In interviews, most people with HIV-associated KS (n=22) identified receiving the transport stipend as a key enabler in their access to treatment, while two people with HIV-associated KS did not mention the transport fare.

> ***I: Out of all we have talked about which one gives you hope of healing KS?***
>
> ***R: The transport fare gave me hope. (CHU 1046, Patient Interview)***

> ***It [transport stipend] has helped me 110 percent because without it, I do not think I would have survived. (ELD 1093, Patient Interview)***

The stipend amount provided seemed to vary with a few people with HIV-associated KS raising the issue of some people receiving more money than others, and suggested that funds be distributed equitably. One person reported that some people did not receive transport assistance for all of their visits:

> ***R: … one of us had already finished the first to six rounds and the disease recurred again…[The peer mentor] used to be giving [Transport fare]…they were not told that the transport is not available so that day when they came they were shocked***

In interviews, navigation team members were able to provide detail regarding the allocation of the transport stipend. People in very remote areas received a greater transport stipend than those patients living closer to the facility. The expectation and hope was that as patients improved with chemotherapy, they would be able to cover their own transport costs.

> ***And then we also have transport reimbursement for patients who have come for their chemo visit. And they get three reimburse[ments] asking for six cycles of chemo, with the expectation that … if there are patients who need more than six cycles, we hope that by the time they get in their sixth cycle, they should be well enough to try and get some money to come for chemo. So just to help the patients to complete their chemotherapy treatment. And that has worked so well, (RCAPRO7001, Navigation Team Interview)***

##### Component Importance

While many patients identified the financial assistance (health insurance stipend and transport stipend) as the most important navigation components. Overall, all aspects of the strategy were considered important.

> ***Everything was important. When you have the transport [reimbursement] and you do not have your papers [NHIF] filled [in], you will not receive treatment…And when you fill the papers (NHIF) without seeing the doctor, you will not get the medication. Everything [referring to navigation components] was important to the patient. (ELD 1093, Patient Interview)***

A strong emerging theme identified in many interviews was the value of connection with the navigators and the value of follow up calls to see how patients were doing between treatment appointments.

> ***Yes, it has appealed to me. Not because bus fare is provided or accommodation, it is because of the relationship between me and the navigators. They are caring, they do follow-ups. That’s the most important thing in this journey. (ELD 1080, Patient Interview)***

All health care workers (n=14) reported that the navigation strategy was important to their facility and most felt it should be integrated into routine care. Many expressed the need to expand the navigation strategy to all oncology patients.

> ***..[the] navigators …compliment whatever the hospital is doing… their absence will be felt…I agree that this [navigation program] should be integrated with our…with our processes…for continuity of treatment. (ELDPRO1009, Health care worker Interview)***

> ***Yes, it [navigation program] should be part of the program, it should be part of the institution, the institution actually has to buy this idea because this is what actually our patients …general oncology patients require and if anything the whole institution needs navigators. (ELDPRO1012, Health care worker Interview)***

One health care worker explained why the strategy was important to both patients and facility staff:

> ***because now if you run it for a short time, those patients will really suffer. Because…I cannot assist the patient… I’ll be dealing with the pharmacy. The records guys can’t assist these patient, I’ve told the patient to look for a file in room 10 C… we are understaffed. You see, so I cannot leave the pharmacy and go and look for the file for the patient or direct a patient to NHIF or social work …I’ll be disadvantaging the other patients who need my service in the pharmacy, them [navigators] geĖng a permanent job will be really… helpful to us, not only the patient, but to us also. (ELDPRO1011, Health care worker Interview)***

Health care workers felt that most of the components of the navigation strategy were sustainable apart from the transport and NHIF stipends. This was reaffirmed by navigation team member interviews where they raised other program’s approaches to sustaining financial assistance:

> ***But then this aspect of …when …money comes in, that becomes a challenge. But I think, I think there are ways to get around that, you know… [other programs] they give them start-ups for small businesses, it could be just one bag of charcoal. So they help you buy one bag of charcoal, they tell you, they talk to you about you know, business, and then …you don’t have to sell that charcoal, you know, your daughter, your son, your husband, your relative can sell the charcoal on your behalf, so that you can raise money to come to clinic.( RCAPRO7001, Navigation Team Interview)***

## Discussion

In sub-Saharan Africa HIV-associated KS continues to be a common and deadly cancer, and despite known evidence-based treatments many people with newly diagnosed HIV-associated KS, who qualify for chemotherapy do not get it. ^1^ ^2^ ^7^ A multicomponent navigation strategy to improve chemotherapy engagement for HIV-associated KS was acceptable and appropriate among people with HIV-associated KS in a mixed-methods analysis and among healthcare workers and navigation team members in a qualitative analysis.

The qualitative data provided a more nuanced and detailed story, where KS patients described which of the six components they felt were most impactful for their cancer journey, and while many people with HIV-associated KS identified the importance of the financial components, others emphasized that all of the components together are what make the strategy helpful and the importance of social support and personal relationships with the navigator and/or peer mentor. Similar themes were echoed among healthcare workers and navigation team members who described the importance of all the components of the navigation strategy, while also identifying challenges in sustaining the financial components of the navigation strategy. Interestingly, even without the financial components, all healthcare workers felt that the multicomponent navigation strategy should be integrated into routine care.

By using implementation science methods and frameworks we were able to clearly define and assess many process measures for this implementation strategy. We found that fidelity to the subcomponents of the implementation strategy as it was designed was variable. Specifically, fidelity was relatively high for certain sub-components of peer mentorship, health insurance assistance, education, and physical navigation. In contrast, fidelity to receiving the transportation stipend for all oncology and chemotherapy visits and receiving the health insurance enrollment stipend were low with only 2.2% and 28% respectively.

In semi-structured interviews, this variability in fidelity was reflected in the experiences of people with KS participating in the navigation strategy. In particular, people with HIV-associated KS commented on variability in appointment reminders, the transportation stipend, and the health insurance enrollment stipend. Interviews with navigation team members identified that using a paper system for tracking patients and their upcoming appointment reminders was error prone and led to some patients not receiving appointment reminders. Navigation team interviews also offered insights into variability in the fidelity to the transportation and health insurance stipends as a by-product of real-world financial constraints to which the navigation team was adapting in real-time. The funding that supported people with HIV-associated KS for transportation, health insurance, and accommodations was principally from donors and there were times when funding was low or unavailable. The team developed a system for rationing the funding, trying to ensure that they were only giving the funding to those with the greatest need. They described struggling to determine the needs of patients and who would receive funding. Because most people work in informal economies where they do not receive any specific documentation of their income, income cut-offs were not feasible, and self-reported income was not always reliable as many do not keep detailed records of their earnings. The struggles with fidelity to the financial components of the strategy bring up questions around the sustainability of the financial components of this strategy and the tension between these being the components that many people with HIV-associated KS described as the most important, yet the most difficult to administer equitably and sustain.

There is limited literature describing evaluation of fidelity to navigation. One study described a detailed evaluation to a navigation strategy to improve the care of people newly diagnosed with schizophrenia.^19^ While they found that most sites in their study had at least basic levels of fidelity, similar to our study there was variability to the components of the “NAVIGATE program,” with highest fidelity to the structure and staffing of the program, and lowest to the “supported employment and education” component of the “NAVIGATE program.”^19^ Similar to our findings the authors concluded that lack of funding and time limitations were important barriers to implementation with fidelity.^19, 20^ Studies of fidelity to implementation strategies are limited in sub-Saharan Africa, but one study evaluating the fidelity to assisted-partner notification for HIV, found that despite the programs overall success, there was variability in the adherence to the details of this complex intervention.^21^ While the program successfully traced the sexual partners of individuals with newly diagnosed HIV, there was variability in how it was done which may be explained by adapting to local realities including weather and reliability of telephone contact information to maximize their likelihood of success.^21^

In our study, all people with newly diagnosed HIV-associated KS who did not participate in the navigation strategy declined to participate in the in-depth interviews. This limits our ability to draw conclusions about why some people with newly diagnosed KS decided not to participate. The majority of people with newly diagnosed HIV-associated KS who did not participate were from a study site that is located 3-4 hours from the AMPATH affiliated cancer center where the navigation strategy was implemented. The inconvenience of long travel times, despite the possibility of a transportation stipend, may have led to lack of participation. Similarly, the financial incentives in the navigation strategy may have led KS patients to speak positively of the strategy in in-depth interviews. We attempted to mitigate this by having interviewers from outside of the KS Center of Excellence of patient in-depth interviews.

In summary, a multicomponent navigation strategy designed to improve chemotherapy engagement for HIV-associated KS was both acceptable and appropriate. Variation in fidelity to the individual components of the intervention suggests areas for future adaptation to ensure sustainability and context appropriateness during integration into the health system and future scale-up.

## Data Availability

All data produced in the present study are available upon reasonable request to the authors.

## Notes

### Competing Interest Statement

The authors have declared no competing interest.

### Funding Statement

This study was funded by the National Institute of Allergy and Infectious Diseases (NIAID), the National Cancer Institute (NCI), and the Fogarty International Center in accordance with the regulatory requirements of the National Institutes of Health under Award Numbers U54 CA190153, U54 CA25457, U54 CA254571-02S1, K23 AI136579, K24 AI141036, and D43 TW009345-09S7 awarded to the Northern Pacific Global Health Fellows Program.

### Author Declarations

The Institutional Review Board of the University of California San Francisco gave ethical approval for this work.

The Institutional Research and Ethics Committee of Moi University gave ethical approval for this work.

